# Preterm premature rupture of membranes before 32 weeks of gestation: Management and pregnancy outcomes

**DOI:** 10.64898/2026.01.14.26344131

**Authors:** Tharwa Ben Necib, Amel Boumaiza, Meriem Garci, Mehdi Makni, Ameni Abdeljabbar, Abir Chaouachi, Olfa Slimani, Nabil Mathlouthi, Cyrine Belghith

## Abstract

Preterm premature rupture of membranes (PPROM) is a major obstetric complication, particularly at early gestational ages. This retrospective, bicentric study (2019–2023) analyzed the short-term outcomes of pregnancies complicated by PPROM between 22 and 32 weeks of gestation under expectant management.

Patients were divided into two groups: G1 (22–26 weeks) and G2 (26–32 weeks). The mean maternal age was 32 years, with a history of PPROM and preterm birth in 26% and 29% of cases, respectively. Antenatal corticosteroids were administered to all patients in G2 and to 28% of those in G1 (p<0.001). Chorioamnionitis was observed in 48% of patients in G1 and 37% in G2 (p=0.183).

The mean gestational age at delivery was 28.03 weeks, with an overall neonatal survival rate of 26% in G1 and 88% in G2 (p<0.001), with survival starting from 25 weeks. Hyaline membrane disease was more frequent in G1 (83% vs. 43%, p<0.001), as was intraventricular hemorrhage without ventricular dilation (70% vs. 27%, p=0.003). Free hyperbilirubinemia was more common in G2 (70% vs. 47%, p=0.002).

The analysis of factors influencing neonatal prognosis highlighted the gestational age at rupture, gestational age at delivery, birth weight, Apgar score, and the amount of amniotic fluid as key determinants.

In conclusion, these factors play a critical role in neonatal prognosis in cases of PPROM.

## INTRODUCTION

Preterm premature rupture of membranes (PPROM) is the spontaneous rupture of fetal membranes before the onset of labor. After 37 weeks of gestation, the main risk is infection, whereas before term, PPROM exposes the fetus to both infectious risks and prematurity. PPROM affects 2–3% of pregnancies, with an incidence below 1% before 34 weeks of gestation [1,2]. Its management is complex due to the lack of consensus regarding fetal viability. Neonatal resuscitation is rare between 22 and 23 weeks of gestation, a gray zone for which thresholds vary by country [2]. A Tunisian study demonstrated that neonatal survival begins at 26 weeks of gestation, with improved outcomes from 27 weeks onward [3].

It is crucial to better understand the management of PPROM between 22 and 32 weeks of gestation and to evaluate short-term neonatal prognosis.

## METHODS

### Study Design

A retrospective, descriptive, and analytical study was conducted to describe therapeutic management and analyze short-term neonatal outcomes.

### Study Setting and Population

A bicentric study was carried out in two maternity units: the “A” Department of Obstetrics and Gynecology at Charles Nicolle University Hospital Center and the Maternity Department of Mongi Slim University Hospital Center, La Marsa, over a period of four years and four months (January 2019 – April 2023).

- **Inclusion criteria**: Patients presenting premature rupture of membranes (PROM) between 22 and 32 weeks of gestation, with an accepted expectant management approach.
- **Exclusion criteria:** Gestational age (GA) at PROM <22 weeks or >32 weeks, multiple pregnancies, delivery occurring within 24 hours after PROM, or contraindication to expectant management (e.g., chorioamnionitis, anhydramnios).

Patients were analyzed globally and then divided into two groups according to GA at the time of PROM:

- Group 1 (G1): 22–26 Week GA
- Group 2 (G2): 26–32 Week GA

### Variables Studied

Socio-demographic, clinical, biological, and ultrasound characteristics; management modalities; delivery details; short-term neonatal complications; and maternal and neonatal factors influencing neonatal outcomes.

### Data Collection

Information was collected from emergencies, hospitalization, and neonatal unit records.

#### Sample Size

A total of 95 patients was included, with 46 in Group 1 and 49 in Group 2.

### Data Analysis

Statistical analyses were performed using SPSS version 25.0. Qualitative data were presented as numbers and percentages, and quantitative data as means, medians, and standard deviations. Analytical comparisons were made using the student’s t-Test and ANOVA. The accepted type I error was 5%, and significance was set at p<0.05.

#### Ethical Considerations

This study was conducted in accordance with ethical principles. Patient anonymity and data confidentiality were strictly maintained.

## RESULTS

PPROM affected 1.2% of the pregnancies studied. The mean maternal age was 32 years (range 21–41). Seventy-six percent of patients were from a low socioeconomic background, and 16% had higher education. Hypertension, a history of preterm delivery, and a previous PPROM were present in less than 50% of cases.

The diagnosis was clinical in 81% of cases, with an Amnisure test performed in 19%. Leukocytosis and a positive C-reactive protein (CRP) were observed in 8% and 21% of cases, respectively. Urine culture (ECBU) and vaginal swabs were positive in 68% and 28% of cases.

At admission, ultrasound revealed oligohydramnios in 60% of cases. All patients received antibiotic therapy. **Antenatal corticosteroid therapy** for lung maturation was administered to all patients in Group 2 and to 28% in Group 1 (p<0.001). Neurological maturation therapy was also more frequent in Group 2. No significant difference was observed regarding tocolysis (Table 1).

**Table 1.** Therapeutic Management in the Two Study Groups.

|  | N/95<br>(%) | Group G1<br>N=46 (%) | Group G2<br>N=49 (%) | P |
| --- | --- | --- | --- | --- |
| Antenatal corticosteroid | 62 (65) | 13(28) | 49 (100) | <0,001 |
| Antenatal neuroprotection | 53 (56) | 12 (26) | 41 (83) | <0,001 |
| Tocolysis | 28 (29) | 10 (22) | 18 (37) | 0.084 |

*N : nombre, G1 : Group 1, G2 : Group 2, P : Statistically significant if $p < 0.05$*

The mean latency period was 7 days (range 3–28 days), with 14 days for Group 1 and 7 days for Group 2.

The mean gestational age at delivery was 28.03 weeks (range 23–34), with 8 patients delivering after 32 weeks. Labor was predominantly spontaneous in Group 1 (63% vs. 41%) and induced in Group 2 (59% vs. 37%), with a significant difference. Chorioamnionitis was the main maternal complication (42%), with no significant difference between groups (p=0.183).

Based on neonatal outcomes, four categories were identified (Table 2). Overall survival was 88% in Group 2 and 26% in Group 1.

**Table 2.** Neonatal Outcomes in the Two Study Groups.

| **Neonatal Outcomes** | N/95<br>(%) | Group G1<br>N/46<br>(%) | Group G2<br>N/49<br>(%) |
| --- | --- | --- | --- |
| Stillbirth | 4 (4) | 4 (9) | 0 (0) |
| Death in delivery room | 6 (6) | 5 (11) | 1 (2) |
| Death in neonatal unit | 30 (32) | 25 (54) | 5 (10) |
| Discharged alive from neonatal unit | 55 (58) | 12(26) | 43 (88) |
| **Perinatal and neonatal mortality** |  |  |  |
| Survived | 55 (58) | 12 (26) | 43 (88) |
| Died | 40 (42) | 34 (74) | 6 (12) |

*N: number, G1: Group 1, G2: Group 2*

Respiratory complications occurred in 67% of neonates, with mechanical ventilation more frequent in Group 1 (83% vs. 43%, p<0.001). Neurological complications were observed in 33% of cases, with intraventricular hemorrhage without dilation more frequent in Group 1 (70% vs. 27%, p=0.003). Early neonatal infections affected 49 neonates (14 in Group 1, 35 in Group 2, p=0.039). Metabolic complications occurred in 93% of cases, with unconjugated hyperbilirubinemia more frequent in Group 2 (70% vs. 47%, p=0.002).

In our series, the analysis of **maternal factors** associated with neonatal outcomes in PPROM identified **gestational age at rupture and at delivery** as the main prognostic factors (Table 3). The mean gestational age at delivery for neonates who died in the neonatal unit was 25 weeks, indicating that neonatal survival was observed from this threshold (Table 3).

Analysis of **neonatal factors** showed that birth weight and Apgar score significantly influenced neonatal prognosis, with higher values among surviving neonates. Additionally, the amount of amniotic fluid at delivery was a key prognostic factor. Among surviving neonates, 53 had oligohydramnios and only 2 had anhydramnios, whereas deaths in the neonatal unit mainly involved neonates with anhydramnios (N=21) and 9 cases with oligohydramnios.

## DISCUSSION

Preterm premature rupture of membranes (PPROM) is a major obstetric complication. In our study, maternal medical and obstetric histories (hypertension, gestational diabetes, preterm delivery, previous PPROM) were present in less than 50% of patients. In contrast, a 2024 meta-analysis identified these as major risk factors, also including recurrent infections [4]. Our ultrasound findings are consistent with a 2020 prospective study on PROM between 23 and 33 weeks of gestation, reporting oligohydramnios in 56.3% of cases and normal amniotic fluid volume in 43.7% [5]. Literature indicates that after PROM, 50–60% of women initially maintain a normal amniotic fluid volume [1,5].

### Management

No outpatient management was implemented in our study. All patients received antibiotics upon admission. Antenatal corticosteroid therapy for lung maturation was administered to all patients in Group 2 and to 28% in Group 1 (p<0.001). Recommendations regarding antenatal corticosteroids vary: the CNGOF recommends initiation at the threshold of neonatal resuscitation, while other societies suggest starting as early as 24 weeks of gestation [2,6,7].

Neurological maturation therapy was prescribed more frequently in Group 2 than in Group 1, with a statistically significant difference. In contrast, no significant difference was observed in tocolysis between the groups.

### Delivery Modalities

Latency was longer in Group 1, consistent with the literature, which shows an inverse relationship between latency duration and gestational age at PROM [8]. The mean gestational age at delivery was 28.03 weeks. Expectant management was maintained for 37 weeks, in line with international recommendations, in the absence of indications for fetal extraction [9,10]. Labor was spontaneous in 52% of cases. Literature reports that 50% of deliveries after PPROM occur spontaneously within 5 days of membrane rupture [11].

Regarding complications, chorioamnionitis was observed more frequently in Group 1, though the difference was not statistically significant. A 2015 retrospective cohort study reported a chorioamnionitis incidence of 57.6% in pregnancies complicated by PPROM [12].

### Neonatal Survival and Mortality

Neonatal survival was 26% in Group 1 and 88% in Group 2 (p<0.001), confirming improved outcomes with increasing gestational age, in agreement with the literature [13].

### Short-term Neonatal Morbidity

PPROM is identified in the literature as a leading cause of prematurity, responsible for 24–42% of preterm births [14]. In our study, all neonates were preterm, with a maximum gestational age at delivery of 34 weeks. Respiratory complications were dominated by hyaline membrane disease (HMD), present in 83% of neonates in Group 1 versus 43% in Group 2 (p<0.001). Yadav et al. [15] reported an HMD incidence of 98% before 24 weeks, decreasing to 5% at 34 weeks and <1% at 37 weeks, confirming the inverse relationship between HMD and gestational age at delivery.

Intraventricular hemorrhage without dilation was observed in 70% of Group 1 and 27% of Group 2, with a significant difference (p=0.003). These results corroborate the literature, which shows that the incidence of IVH is inversely proportional to gestational age at delivery [16].

Infectious complications, such as early-onset neonatal sepsis, result from maternal-fetal transmission via transplacental, ascending, or peripartum routes, with Group B Streptococcus and Escherichia coli being the main causative organisms. Incidence is higher in low-birth-weight neonates and those born before 28 weeks [17]. Bohiltea et al. reported a decreased incidence of neonatal sepsis with advancing gestational age [18]. In our study, neonatal infections were more frequent in Group 2, which could be due to the smaller sample size.

### Maternal and Neonatal Factors Associated with Neonatal Outcomes

Gestational age at rupture and at delivery were the main prognostic factors for neonatal outcomes. Improved outcomes were observed with later delivery after PPROM, with gestational age at delivery being more decisive than at rupture [14]. Birth weight and Apgar score also predicted neonatal outcomes, consistent with the literature [19].

The impact of amniotic fluid volume on neonatal outcomes has been studied. A 2023 retrospective study in a level III neonatal intensive care unit demonstrated that oligohydramnios is significantly associated with increased perinatal and neonatal mortality, mainly due to the higher risk of pulmonary hypoplasia [20].

### Strengths and Limitations

The strengths of this study include its bicentric design and comparative analysis between groups, allowing identification of factors influencing neonatal outcomes. Limitations include its retrospective nature and focus only on short-term neonatal prognosis.

## Conclusion

Preterm premature rupture of membranes is a major obstetric complication, with management complexity depending on gestational age and neonatal resuscitation capabilities. Neonatal complications are primarily related to prematurity, and prognosis depends on gestational age at membrane rupture, gestational age at delivery, birth weight, Apgar score, and amniotic fluid volume. Further studies are needed to assess medium- and long-term neonatal outcomes.

## Data Availability

All data produced in the present work are contained in the manuscript

